# Interpretable multivariate survival models: Improving predictions for conversion from mild cognitive impairment to Alzheimer’s disease (AD) via data fusion and machine learning

**DOI:** 10.1101/2025.03.11.25323801

**Authors:** Diana Rosales-Gurmendi, Gerardo Fumagal-González, Jorge Orozco-Sanchez, Joshua Farber, Victor Treviño, Emmanuel Martinez-Ledesma, Antonio Martinez-Torteya, Fabiola Rosales-Gurmendi, Jose Tamez-Peña, Alzheimer’s Disease Neuroimaging Initiative

## Abstract

Accurately predicting which individuals with mild cognitive impairment (MCI) will progress to Alzheimer’s disease (AD) can improve patient care. This study examines the role of quantitative MRI (qMRI), cognitive evaluations, apolipoprotein *ε*4 (*APOE ε*4), and cerebrospinal fluid (CSF) biomarkers in Cox survival models to predict progression from MCI to AD. Data from 564 participants in the ADNI study, who transitioned from MCI to AD, were analyzed. The data set included 330 features encompassing qMRI, cognitive assessments, CSF biomarkers, and *APOE ε*4 status. Advanced machine learning (ML) methods were applied to evaluate the importance of these data sources, select relevant features, and develop interpretable Cox survival models within a cross-validation framework. The top optimized model achieved a sensitivity of 0.69, 95% CI [0.63, 0.76], and a specificity of 0.87, 95% CI [0.83, 0.90], and used all data sources. The results demonstrated that combining qMRI features with cognitive assessments, CSF biomarkers, and *APOE ε*4 status, analyzed using the BSWiMS model, resulted in a substantial improvement in the ability to predict progression from MCI to AD, achieving 81% precission and 87% specificity. These results exceed those obtained with other models evaluated. Finally, biomarker analysis showed that cognitive scores are the most relevant features to predict conversion, followed by CSF and qMRI biomarkers. These findings highlight the value of integrating multiple data sources in highly interpretable Cox survival models for the early identification of individuals at risk for Alzheimer’s disease.

## Introduction

Alzheimer’s disease (AD) is one of the most common cognitive disorders in old age [1]. The development of effective treatments or disease-modifying therapies is hampered by the complexity of aging and the lack of a clear understanding of the etiology and pathogenesis of AD [2]. The diagnosis of AD in the early stages of the disease is complex. The most accurate diagnostic test for AD requires a histopathological evaluation of brain tissue by autopsy or biopsy [3]. Without a biopsy, the diagnosis of a normal patient is defined as possible or probable AD according to patient reports, cognitive observation, and symptomatology [4]. Hence, understanding the process and each of the stages of AD is essential to developing effective treatments. It is noteworthy that mild cognitive impairment (MCI) is regarded as a transitional stage between normal aging and AD [5]. Therefore, in the context of the conversion of mild cognitive impairment (MCI) to AD, some biomarkers have been identified in the AD literature. The beta-amyloid and tau proteins from cerebrospinal fluid (CSF) [6, 7], the scores in cognitive assessments [8], and the polymorphism of the fourth allele of the apolipoprotein E (*APOE ε*4) [9] have been recognized as validated risk factors for the conversion. On the other hand, some studies have shown that clinical information and imaging biomarkers can be used to predict patients who will undergo conversion from those who will not [10]. Imaging biomarkers related to AD have been found in PET and MRI and have a clear association with the evolution and presence of AD [11] and with the conversion of MCI to AD [4]. However, the impact of imaging biomarkers to determine the conversion rate between MCI to AD, as well as the relationship and the importance of the MRI features in survival models, have not been fully studied.

Studies indicate that CSF biomarkers are increasingly being used to support the diagnosis of Alzheimer’s disease, especially to determine the difference between AD from non-Alzheimer’s disease dementia [12, 13, 14]. Different biomarkers can be extracted from CSF. The most established in the usual clinical practice include beta-amyloid 1-42, total tau (t-tau), and phosphorylated tau in threonine 181 (p-tau) [12]. All of them have been previously described in AD pathogenesis. Despite their role in AD pathogenesis the cerebrospinal fluid biomarkers have an important utility for AX-42/X-40 and T-tau/A1-42 ratios, but limited specificity for distinguishing AD from DLB and PNFA [14, 15].

Alternatively, the symptomatic AD process alters brain function; henceforth, cognitive assessments are usually the starting point for the brain health plan and are recommended for all patients who want to be screened for AD [16, 17]. These assessments evaluate some important areas of brain function, memory and reasoning capacity, concentration, processing speed, and language. Depending on the country, language, or type of test, several experts have developed and validated standard tests such as those used in this work. Some authors have updated and proposed changes in the normal cognitive assessments to better classify or predict the conversion in patients, [18] and others proposed the combination of patterns of brain atrophy and cognitive assessment scores, the latter offering the highest predictive power for the conversion [19].

Other tests available for the understanding of the AD process are magnetic resonance imaging (MRI) and positron-emitting tomography (PET). These imaging modalities can visualize direct changes in brain structure, function, sugar metabolism, and AD build-up [20, 21]. Therefore, AD-related imaging biomarkers with a clear association on the early development and presence of AD have been discovered in MRI and PET [11]. Furthermore, imaging biomarkers have been associated with the conversion from MCI to AD [4, 22]. Consequently, MRI and PET are commonly used to monitor the progression of the disease and to detect the current stage of neuronal degeneration [23].

Lastly, the onset of AD requires a clear description of its risk factors. Age and gender are established risk factors for developing AD. Older people are at higher risk, while women are more prone than men to developing AD. *APOE ε*4, the polymorphic allele of the apolipoprotein *ε*, is a known genetic risk factor for developing AD [23]. These known factors must be considered when developing screening or staging AD tests. Therefore, researchers have proposed the combination of risk factors, cognitive tests, MRI, and PET features with CSF biomarkers to improve the diagnostic accuracy of AD and to improve the predictive power of MCI to AD conversion [6].

The development of a good screening test requires the identification, characterization, and validation of biomarkers associated with AD and requires a good understanding of their evolution as well as their role in the disease process. Although there is plenty of research describing the association between biomarkers at the MCI stage and probable AD conversion, their discoveries have not effectively evaluated the time required from MCI to AD conversion [23, 24, 25]. This evaluation is important because some biomarkers may be associated with a slow conversion process (low-risk markers) while others with a swift conversion (high-risk markers) [26]. In this context, multivariate Cox regression models are a statistical tool that can be used to screen out low-risk markers vs. high-risk markers. Cox modeling incorporates the time to event in its fitting process and provides estimates of the hazard ratios (HR) of each potential biomarker. This feature of Cox modeling can be used to improve the understanding of biomarkers associated with the AD process. In addition, Cox proportional hazard has been widely used in survival studies [27].

The challenge of Cox modeling in biomarker discovery is that thousands of potential biomarkers can be associated with the risk of conversion. Traditional approaches use hypothesis-based feature selection; thus, findings have been limited to a small set of biomarkers [28]. Alternatives to the traditional approach are subset selection and regularization. Subset selection is based on computer algorithms that attempt to identify the best set of features associated with the disease process. Regularization methods use all the available features to estimate the total multivariate risk, and they solve the ill-posed problem by adding heuristic constraints. Statistical learning (SL) and machine learning (ML) strategies provide efficient and highly competitive regularization and subset selection methods. Embedded SL approaches like L1 regularization via LASSO or L2 regularization through RIDGE allow the exploration of multivariate models composed of hundreds of features [29]. L1 regularization via the LASSO also allows subset selection [30]. Bootstrap Step-Wise Model Selection (BSWiMS), golden section primal-dual active set (GPDAS), and sequential primal-dual active set (SPDAS) are other subset selection methods readily available to researchers [30, 31]. Finally, researchers usually rely on traditional statistical methods controlled for false discovery rate (FDR) for feature selection (FS) [32, 33].

The wide variety of methods available to researchers can make biomarker discovery a complex effort, especially when there is no clear choice of methodology for building/exploring survival models. To overcome this limitation, we propose a unified approach for the study of Cox models in an ML setting. The approach is based on repeated cross-validating ML methods using the same training-testing sets across all the methods. The implementation evaluates LASSO, RIDGE, BSWiMS, GPDAS, SPDAS and FDR-controlled univariate filtering for building suitable survival models [34]. Thus, the result of the approach is a fair method comparison and a comprehensive evaluation of the role of each potential biomarker inside a Cox survival model. The primary goal of this paper is to provide a strict comparison of Cox models constructed from multivariate-multi-data sources (imaging, risk factors, CSF, and cognitive assessments) that describe the MCI to AD conversion risk for each subject in a unified approach. Additionally, this study aims to highlight the relevance of the multiple biomarkers that are present in the conversion process of MCI to AD. Furthermore, a key objective is to provide a unique consensus-based Cox model that can be used to accurately predict the chances of conversion of MCI-diagnosed subjects. Subsequent sections present the data preparation, the utility of the unified approach for the comparison of ML models, and the role of the top biomarkers associated with MCI to AD conversion.

## Materials and methods

Data used in this study was obtained from TADPOLE challenge dataset (https://tadpole.grand-challenge.org), this dataset was made publicly available and anonymized since 16 June 2017, which was derived from the ADNI database (adni.loni.usc.edu). The ADNI was launched in 2003 as a public-private partnership, led by Principal Investigator Michael W. Weiner, MD. ADNI is a cross-sectional and longitudinal follow-up observational study. The main goal of ADNI has been to see how neuroimaging, cognitive tests, fluid and genetic biomarkers can be used together to figure out how MCI and early AD will progress. For up-to-date information, see www.adni-info.org.

The ADNI/TADPOLE challenge datasets considered for this study were “D1—a comprehensive longitudinal data set for training”, and “D2—a comprehensive longitudinal data set on rollover subjects for forecasting”. The challenge included 1,737 individuals from the ADNI database with longitudinal observations. Each subject’s data included the diagnosis status, cognitive assessments, qMRI and PET data, and *APOE ε*4 status. Detailed information regarding the rationale and the features contained in the TADPOLE challenge [33].

### Ethics considerations

As mentioned earlier, data used in this study is publicly available and therefore no additional ethical approval was needed from an ethics committee, however, written informed consent for ADNI participants was obtained by the ADNI in accordance with the local legislation and ADNI requirements. ADNI studies follow Good Clinical Practices guidelines, the Declaration of Helsinki, and United States regulations (U.S. 21 CFR Part 50 and Part 56).

### Subjects

The general eligibility, inclusion, and exclusion criteria used in this study are summarized in Fig 1. From a total of 1,737 individuals with a baseline diagnosis of MCI, who were common among the TADPOLE individuals (30%), 622 of these subjects were further excluded due to not having complete observation baseline data; in that sense, only 1,115 subjects met criteria for inclusion as part of the MCI group recruited for ADNI. Likewise, 551 of these subjects were excluded for having the condition of non-convert (NC) and Alzheimer’s diagnosis (AD). Finally, 564 MCI subjects were included, with 191 subjects converted and 373 non-converted to assess the difference between groups in the study and to evaluate the importance of features.

**Fig 1.**
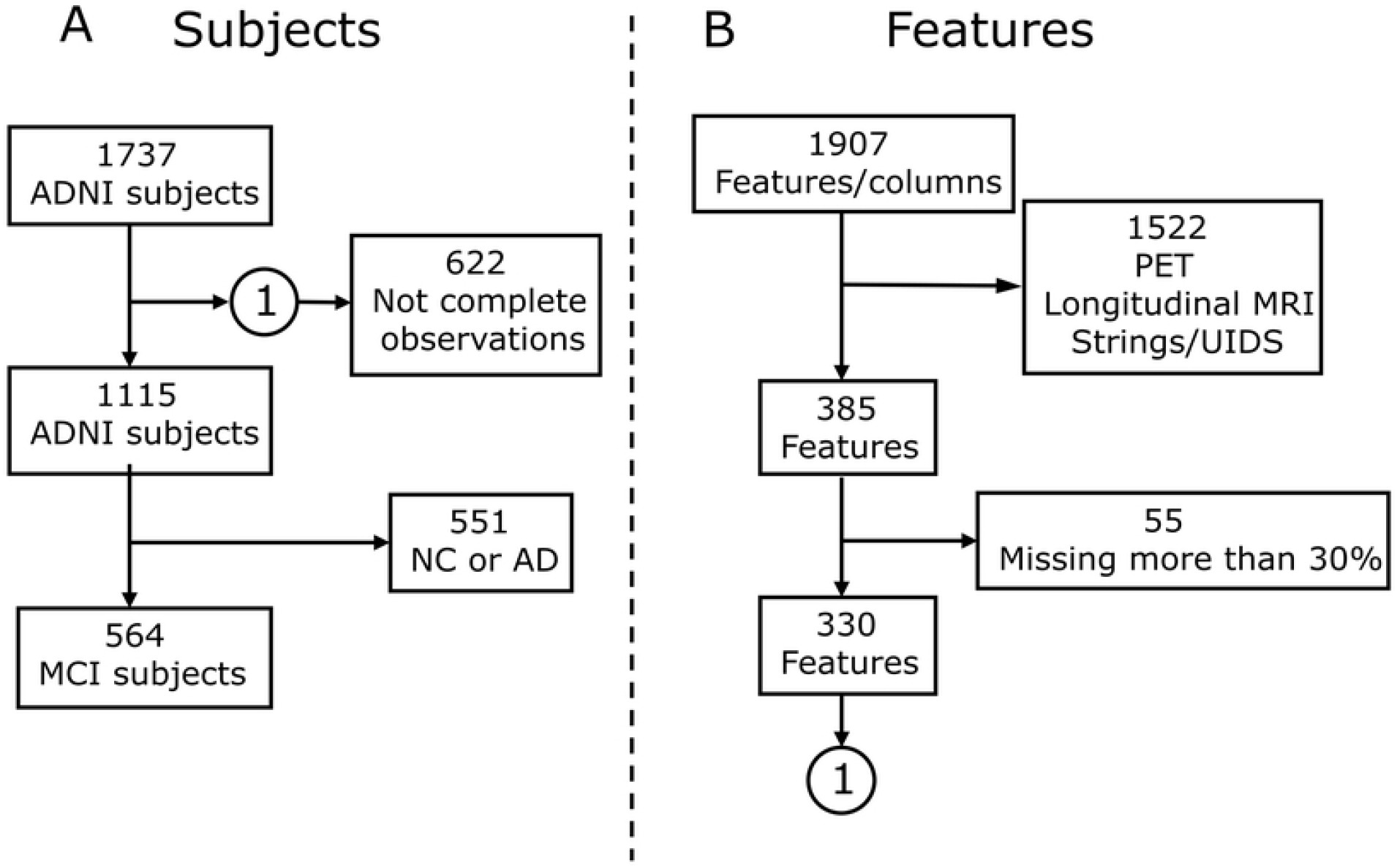
Data inclusion and exclusion diagram: **(A)** Subject selection, a baseline of 1737 subjects from ADNI; 622 subjects with missing data or few observations were excluded and 564 subjects with MCI and complete observations were included. **(B)** Feature selection, a total of 1907 features were identified from ADNI-TADPOLE. Of these, 1522 PET and longitudinal analysis features with Free Surfer were excluded, along with features containing missing data, leaving 330 features per patient.

### Clinical Data

We considered the main features as potential predictors of MCI to dementia conversion in our analysis; therefore we divided the features into three major groups. The features detailing levels of three different proteins, (A*β*_1*™*42_), t-tau, and p-tau, were included and studied in a group labeled as CSF features. These measures were obtained during baseline evaluation at the University of Pennsylvania Medical Center. In addition, we included information from several neuropsychological tests, labeling the entire group of features as cognitive assessment features. Due to the nature of the disease, an ADNI examiner interviewed the patient to determine all the cognitive assessment scores. A complete description of the assessment acquisition is found in the ADNI manual. Likewise, seven assessments were used: Clinical Dementia Rating Sum of Boxes (CDRSB) [34], Alzheimer’s Disease Assessment Scale (ADAS) [35], Mini-Mental State Examination (MMSE) [36], Rey’s Auditory Verbal Learning Test (RAVLT) [37], and Functional Assessment Questionnaire (FAQ) [38]. The 346 qMRI measurements provided by the University of California San Francisco (UCSF) were included and labeled as Radiomic features. Each MRI dataset was post-processed using FreeSurfer v4.3, a processing program with the function of (a) automated model-based reconstruction of the brain’s cortical surface and subcortical structures and (b) morphometric analysis. Finally, sex and *APOE ε*4 status were also included in this group. Table 1 shows the descriptive characteristics of the main features of the dataset.

**Table 1.**
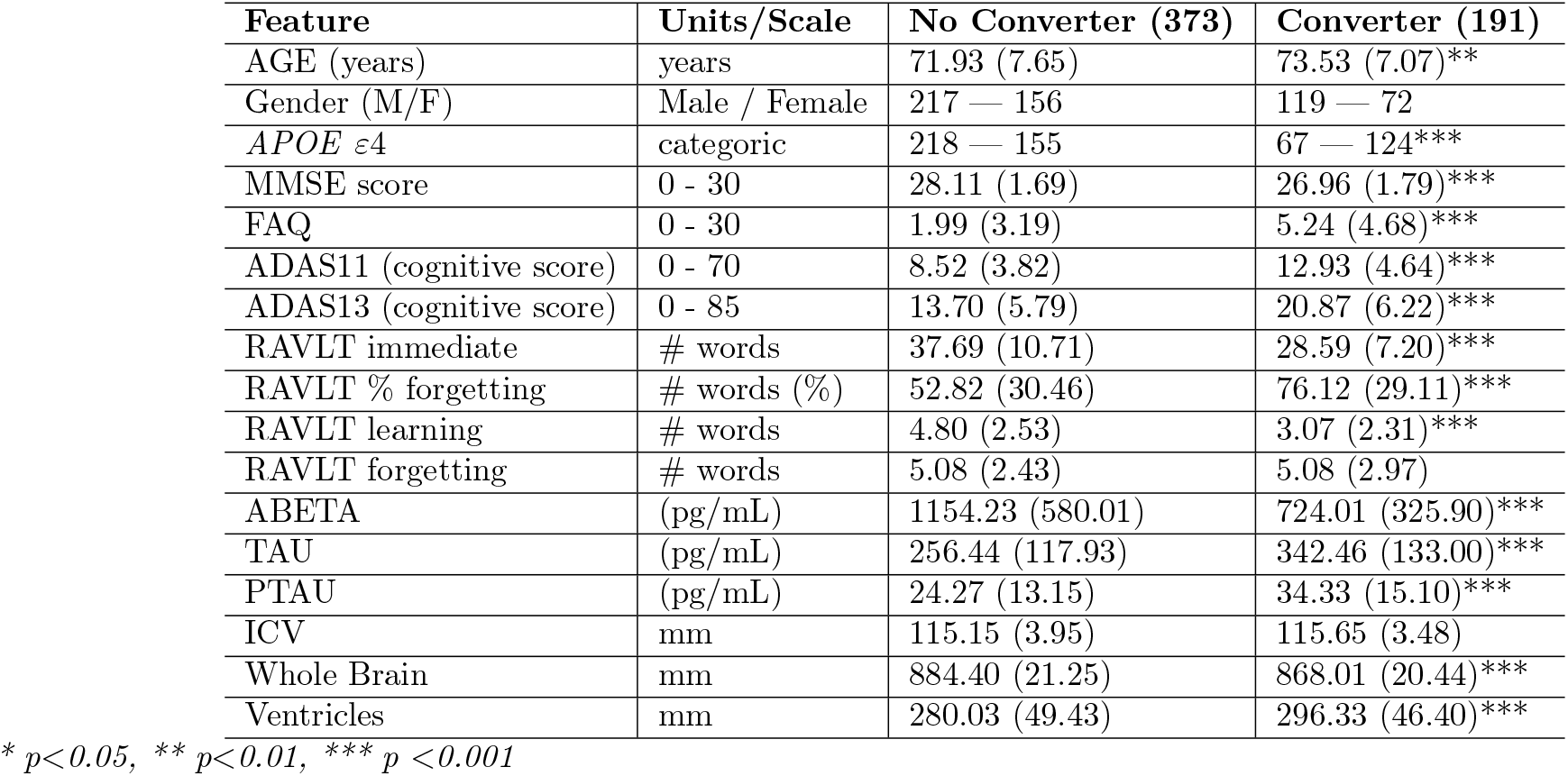
Demographic data with features sectioned by non-converter and converter groups.

Based on the previously mentioned data, the methodology process can be outlined in Fig 2. In which it begins with the inclusion and exclusion of data from the ADNI-TADPOLE set. This is followed by a rigorous preprocessing phase focusing on individuals with MCI, aimed at harmonizing the diverse data sources. Subsequently, the process transitions into a comprehensive evaluation of machine learning models, comparing their predictive performance to determine the most accurate configurations for identify MCI to Alzheimer’s conversion. To complement these analyses, six Cox regression models, adjusted for survival analyses, are compared to evaluate their capacity for predicting progression over time. Each stage of the workflow incorporates specific procedures, such as cross-validation, to minimize inter-cohort bias and ensure the validity and reliability of the results. This integrative approach not only strengthens the reproducibility of findings but also emphasizes their clinical relevance, providing actionable insights into the progression of Alzheimer’s disease.

**Fig 2.**
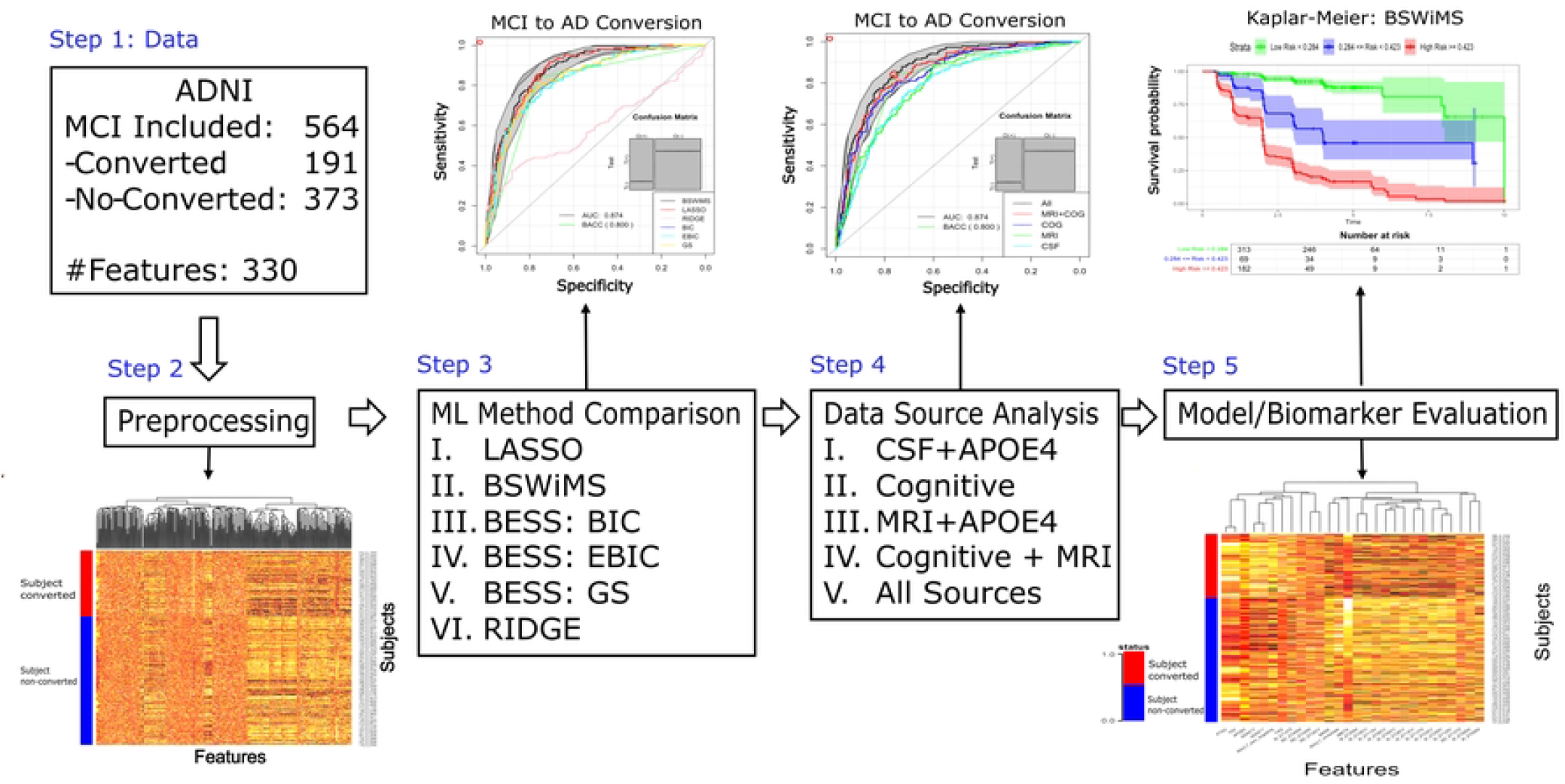
Diagram, the process of analysis of discovered biomarkers, data fusion (CSF: cerebrospinal fluid, MRI: magnetic resonance imaging, Cog: Cognitive, *APOE ε*4 (apolipoprotein E4)) by cross-validation and Machine Learning methods comparison evaluating the performance of 6 Cox models.

### Data conditioning and pre-processing

We computed the time to conversion from the provided data. The event time for subjects that converted consisted of the difference in days between the date of their first AD diagnosis and the baseline date. The event time for stable MCI subjects consisted of the difference in days between the date of the baseline and the date of the last recorded follow-up visit. MCI-stable subjects were labeled as censored. We pre-processed the radiomic features as follows: We compute the cubic root of all qMRI volumes, and the square root of all qMRI areas. All qMRI data was normalized by dividing each one by intracranial volume. After that, the qMRI measures of the left and right sides of the brain were described by the mean, absolute, and relative differences. Hence, these last features can be used to check dementia issues due to brain asymmetry.

### Cox modeling via Machine Learning Subset Selection

To explore the subset of features and its association with the MCI to AD conversion, we used machine learning to train Cox regression models. Cox models explore the relationship between the time of the event and the possible explanatory variables. The model estimates the hazard, *λ*_*i*_, of the subject *i* given the observed feature vector *X*_*i*_ = *{X*_*i*1_, …, *X*_*ip*_*}*, and the unknown baseline hazard *λ*(*t*_0_), i.e given by:

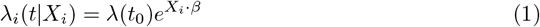

where *β* = {*β*_1_, …, *β*_*p*_} is the vector of coefficients. The fitting is commonly performed by a maximum likelihood estimation method providing the *β* values and relative hazard ratios. Thus, the Cox model provides an estimate of the total hazard or risk of conversion, given the observed features for an individual. Due to the large set of possible qMRI features to be considered in some of the Cox models, ML methods were used to find an optimal set of features and their corresponding coefficients that mimicked the observed rate of conversion.

There are several strategies for building Cox models [29, 39, 40, 41, 42]. In this paper, we evaluated three open-source ML packages. These packages provided us with different strategies for building Cox models. The first ML method was BSWiMS, part of the FRESA.CAD R package [29], is a supervised model-selection method aimed at generating a unique statistical model that predicts a user-specified outcome, in this case, a survival outcome. The statistical model is constructed by bagging a set of Cox models, where each Cox model is composed of a set of model-wise statistically significant features [40]. The second evaluated package was the gmlnet R package that implements the Penalized Cox Regression (CoxNet) algorithm. CoxNet fits the Cox model regularized by L1 or L2 or a mixture of them using the elastic net penalty [43]. We executed CoxNet with their provided 10-fold cross-validation function (cv.glmnet) to determine the optimal weight of the regularization constraint. Furthermore, we ran cv.glmnet with two alpha values: alpha=1 (feature selection with LASSO regularization) and alpha=0 (ridge or L2 regularization). The third evaluated package was the BeSS R package. This package implements the Golden Section Primal-Dual Active Set algorithm (GSPDAS), aiming for the selection of the best set of features of the Cox model. Like CoxNet, BeSS can use different strategies for subset selection. The default configuration uses the GSPDAS algorithm. The second BeSS option is SPDAS based on the Bayesian information criterion (BIC). The third option runs SPDAS algorithm with the EBIC. In summary, we evaluated six different Cox models: BSWiMS, LASSO, RIDGE, BESS:BIC, BESS:EBIC, and BESS:GS.

### Cox model validation and evaluation

The main aim of this paper was the comprehensive evaluation of the Cox models for the prediction of the MCI to AD conversion. To ensure a fair comparison, we employed a repeated holdout cross-validation (RHOCV) approach across all machine learning (ML) strategies. This consistent evaluation framework was applied to each set of features provided by the TADPOLE challenge (Fig 2). The test results of the RHOCV were used to compare and explore the performance of the ML alternatives. The RHOCV strategy is part of the FRESA.CAD R package. The RHOCV method creates multiple sets of training and testing sets. At each interaction, the input data is randomly divided into a training and testing set. The training set is used for the model or feature selection, while the holdout set is predicted by the trained method. Once all the holdout predictions are generated, the test results are evaluated and compared between ML strategies. The RHOCV implementation uses the Survival R package to calculate the final Cox predictions of each model.

The estimated coefficients *β*^*j*^ on each training set *T*_*j*_ were used to get the subject *i* linear predictions 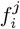 of the holdout set 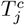 at each repetition:

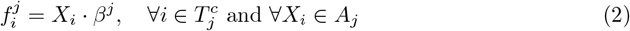

Once the linear predictions were obtained for each repetition, the test results were ensembled by computing the median prediction for each subject as 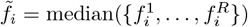. The ensemble prediction was used to divide the subjects into high-risk 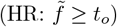 and low-risk 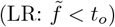 groups, where *t*_*o*_ is the decision threshold. We assumed that censored subjects belong to the low-risk group, while true MCI to AD conversions are in the high-risk group. The receiver operating characteristic (ROC) plots and their area under the curve (AUC) with their corresponding 95% confidence intervals (CI) were computed for the risk prediction using the pROC package [44]. Accuracy (ACC), sensitivity (SEN), and specificity (SPE), describing the ability of the Cox models to predict censored vs. uncensored subjects, were computed based on the number of true positives (TP) and true negatives (TN) given by:

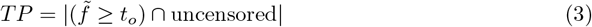

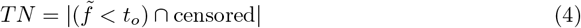

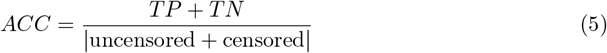

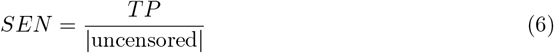

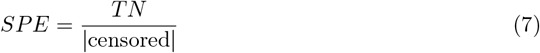

The survival performance was evaluated using the concordance index (c-index). The c-index measures the fraction of all order pairs of subjects *ϵ*_*ij*_ Whose predicted survival times are correctly ordered among all subjects that can be ordered. It can be written as:

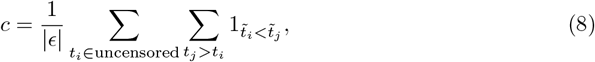

where the indicator function 1_*a<b*_ = 1 if *a < b*, and 0 otherwise, |*ϵ*| is the number of ordered pairs. 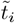 is the median of the predicted survival time and *t*_*i*_ is the actual observed time of the uncensored subject *i*. The values of the c-index range from 0 to 1, where 1 implies a perfect concordance between observed and predicted times.

We evaluated the prediction benefit using decision curve analysis (DCA). The DCA curves indicated a net benefit of the predicted probability of the models. The analysis of the predicted probability was used to estimate the high-risk threshold at 90% specificity and the middle-risk threshold at 80% specificity. The 90% threshold was then used to evaluate the classification performance of the models.

The visualization of the predicted survival groups, high-risk vs. mid-risk and low-risk, was done using Kaplan-Meier (KM) plots of the survminer R package [45]. The statistical significance of the difference between the survival groups was evaluated by the log-rank test [46] given by:

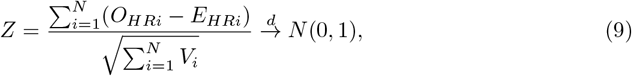

and

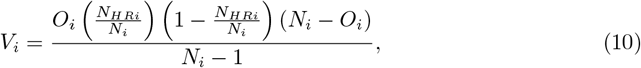

where *O*_*i*_ is the actual number of events, *N*_*i*_ is the number of subjects below rank, and *N*_*HRi*_ is the number of subjects at high risk.

### Feature source and Cox-models

To evaluate the predictive performance of the Cox models, we conducted a series of experiments using the Recursive Hyperparameter Optimization with Cross-Validation (RHOCV) procedure, repeated 50 times for robustness. Each run allocated 70% of the subjects for training and 30% for testing, and the models were constructed by selecting features dynamically in each iteration. This approach enabled the identification of the most frequently selected features across experiments, providing insight into their relative importance. The experiments were designed to assess the impact of different data sources on the models. Five independent experiments were performed: Experiment 1 used CSF and *APOE ε*4 features. Experiment 2 studied six Cognitive Assessment features. Experiment 3 constructed models from the MRI and *APOE ε*4. Experiment 4 used cognitive assessment and MRI. Experiment 5 used all the features from CSF, cognitive assessment, MRI, and *APOE ε*4 as part of the feature set. Each experiment consisted of the independent execution of the RHOCV procedure 50 times. Each run used 70% of the subjects for training while 30% was used for testing. The procedure builds models selecting features in each run; hence the analysis plan included the exploration of the common top features discovered in each experiment.

### Biomarker evaluation via model feature analysis

The last experiment was designed to explore the hazard ratios of the top Cox model features associated with the MCI to AD conversion. First, we set the training (70%) and testing sets (30%). Then we z-standardized all features using the training set to estimate the mean and variance of the continuous features and estimated the z-scores of the testing set. After that, we run the BSWiMS procedure for 20 Cox bootstrap estimations using the training set and evaluating the performance on the testing set. The summary method of the BSWiMS analyzes the hazard ratio of each fitted model and reports their 95% confidence intervals for each one of the top features associated with conversion.

## Results

The preceding section detailed the materials and methods used to construct and evaluate various machine learning (ML) models aimed at predicting the conversion from mild cognitive impairment (MCI) to Alzheimer’s disease (AD). This section presents the findings obtained by applying these models to the TADPOLE dataset.

The objective was to identify the optimal ML model and feature selection method for accurately forecasting the progression from MCI to AD. To achieve this, we assessed the performance of six distinct ML algorithms using metriscs such as the area under the receiver operating characteristic curve (AUC), accuracy, sensitivity, and specificity.

### ML feature subset selection method

The performance of six machine learning (ML) models was evaluated on independent test sets. Table 2 summarizes the results, including ROC AUC, the c-index, the accuracy, the sensitivity, and the specificity of all six evaluated ML methods on the testing sets. Fig 3 shows the ROC curve of all the evaluated ML methods, decision curve analysis (DCA) and the Kaplan-Meier of the BSWiMS, BESS:BIC, and LASSO test results. The BSWiMS model showed a test performance with AUC 0.87, 95% CI [0.85, 0.90], SEN 0.69, 95% CI [0.63, 0.76], and SPE 0.87 at 95% CI [0.83, 0.90]. The LASSO model also showed an equivalent performance, with AUC 0.88 at 95% CI [0.85, 0.90], SEN 0.64 at 95% CI [0.57, 0.71], and SPE 0.87 at 95% CI [0.83, 0.90]. The BSWiMS and LASSO were statistically similar, with the advantage that BSWiMS models’ coefficients are highly interpretable. The BeSS models and RIDGE regression did not produce results as good as the LASSO and BSWiMS. These models were statistically inferior to the BSWiMS methods; furthermore, the DCA analysis indicated an inferior benefit to predict subjects that will convert to AD. The worst-performing method was the RIDGE method, whose ROC AUC was 0.54 at 95% CI [0.49, 0.58]. All models were able to stratify the high-low risk groups with statistical significance.

**Table 2.** ML subset selection method comparison.

| Method | ROC AUC | c-Index | Accuracy | Sensitivity | Specificity |
| --- | --- | --- | --- | --- | --- |
| BSWiMS | 0.87 [0.85,0.90] | 0.85 [0.82,0.87] | 0.81 [0.78,0.84] | 0.69 [0.63,0.76] | 0.87 [0.83,0.90] |
| BESS:BIC | 0.83 [0.80,0.87] | 0.82 [0.79,0.84] | 0.79 [0.75,0.82] | 0.63 [0.56,0.70] | 0.86 [0.83,0.90] |
| BESS:EBIC | 0.83 [0.80,0.87] | 0.82 [0.79,0.84] | 0.79 [0.75,0.82] | 0.63 [0.56,0.70] | 0.86 [0.83,0.90] |
| BESS:GS | 0.84 [0.80,0.87] | 0.82 [0.79,0.85] | 0.79 [0.76,0.82] | 0.63 [0.56,0.70] | 0.87 [0.83,0.90] |
| LASSO | 0.88 [0.85,0.90] | 0.85 [0.82,0.87] | 0.79 [0.75,0.82] | 0.64 [0.57,0.71] | 0.87 [0.83,0.90] |
| RIDGE | 0.54 [0.49,0.58] | 0.57 [0.53,0.61] | 0.56 [0.52,0.60] | 0.48 [0.41,0.55] | 0.61 [0.56,0.66] |

**Fig 3.**
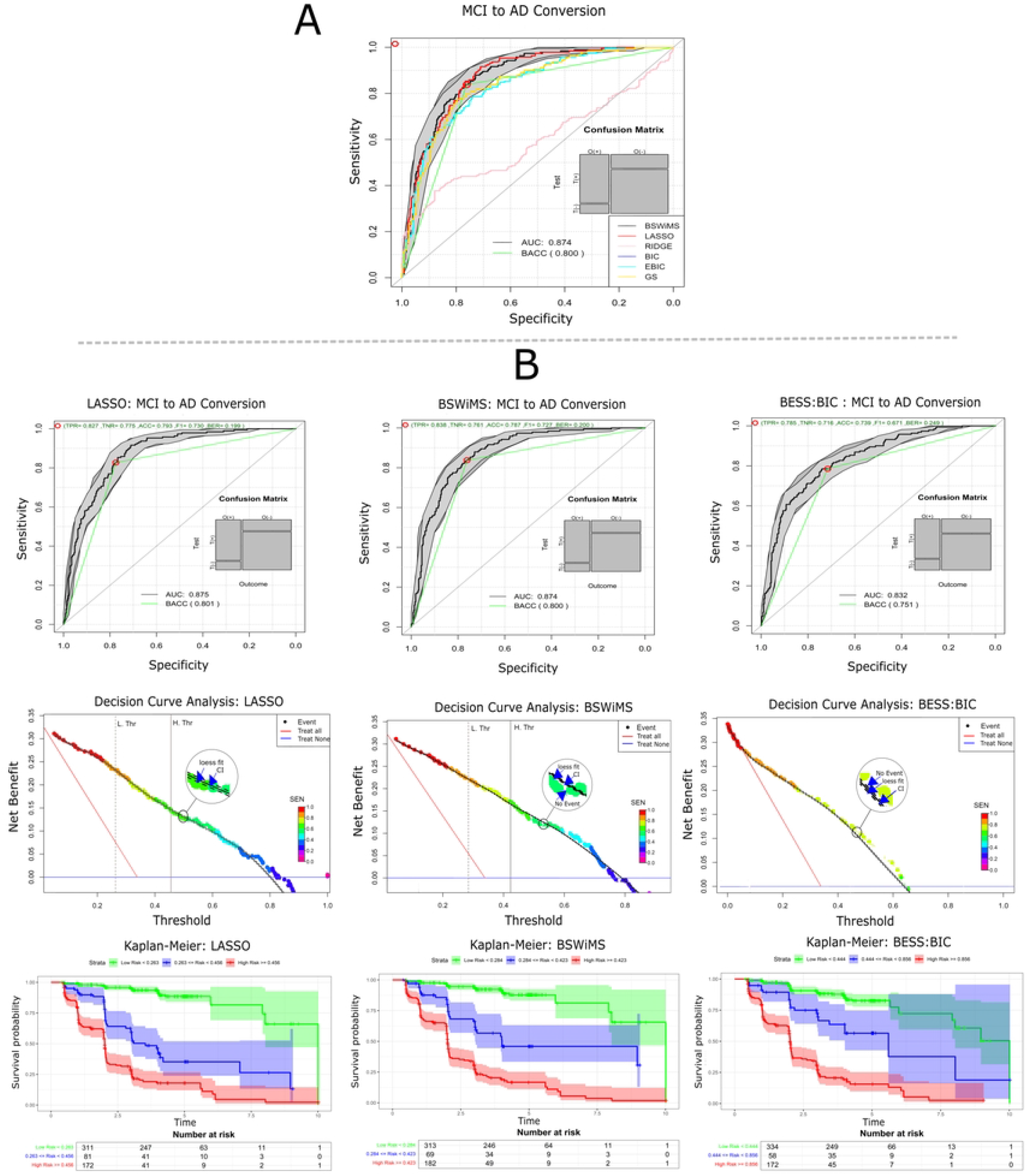
Evaluation and survival model analysis comparison. **(A)** Comparison of all ROC curves of all source-experiments. **(B)** ROC curves, Decision Curve, and Kaplan-Meier of LASSO, BSWiMS, and BESS BIC, respectively.

### Features source and Cox-models

To evaluate the impact of different data sources on model performance, Cox proportional hazards models were trained and evaluated using five distinct sets of features. Table 3 reports the performance metrics of these Cox models, and Fig 4 shows the ROC AUC for all five sources as well as the DCA analysis and Kaplan-Meier of the cognitive and CSF features. The model developed using CSF and qMRI data showed the lowest classification performance, with an AUC of 0.78 at 95% CI [0.74, 0.82] and 0.79 at 95% CI [0.75, 0.83] respectively. The cognitive model showed the best performance of 0.84 at 95% CI [0.81, 0.87] that was statistically superior to MRI and CSF models (p *<*0.001). The models that used multiple data sources were statistically superior to the cognitive-only model. ROC AUC improved to 0.85, 95% CI [0.82, 0.89] for the qMRI-Cognitive model and to 0.87, 95% CI [0.85, 0.90] for the model that used all the data sources. All these models that fused data sources were statistically superior (p *<*0.01) to the models that only used a subset of features. The Kaplan-Meier analysis indicated that the model that used all the data sources (Fig 3) showed subjects classified as low risk had a nice five-year non-conversion rate (87%) when compared to the high-risk group that by year 5 more than 80% of them had converted to AD.

**Table 3.** Source Comparison on BSWiMS models.

| Model | ROC AUC | c-Index | Accuracy | Sensitivity | Specificity |
| --- | --- | --- | --- | --- | --- |
| All | 0.87 [0.85,0.90] | 0.85 [0.82,0.87] | 0.81 [0.78,0.84] | 0.69 [0.63,0.76] | 0.87 [0.83,0.90] |
| Cog + MRI | 0.85 [0.82,0.89] | 0.83 [0.81,0.86] | 0.79 [0.76,0.83] | 0.65 [0.58,0.72] | 0.87 [0.83,0.90] |
| Cog | 0.84 [0.81,0.87] | 0.83 [0.80,0.85] | 0.79 [0.75,0.82] | 0.62 [0.55,0.69] | 0.87 [0.83,0.90] |
| MRI+ *APOE* $\epsilon 4$ | 0.79 [0.75,0.83] | 0.76 [0.73,0.80] | 0.74 [0.70,0.77] | 0.47 [0.40,0.54] | 0.87 [0.84,0.91] |
| CSF+ *APOE* $\epsilon 4$ | 0.78 [0.74,0.82] | 0.73 [0.69,0.76] | 0.72 [0.68,0.76] | 0.40 [0.33,0.47] | 0.88 [0.85,0.92] |

**Fig 4.**
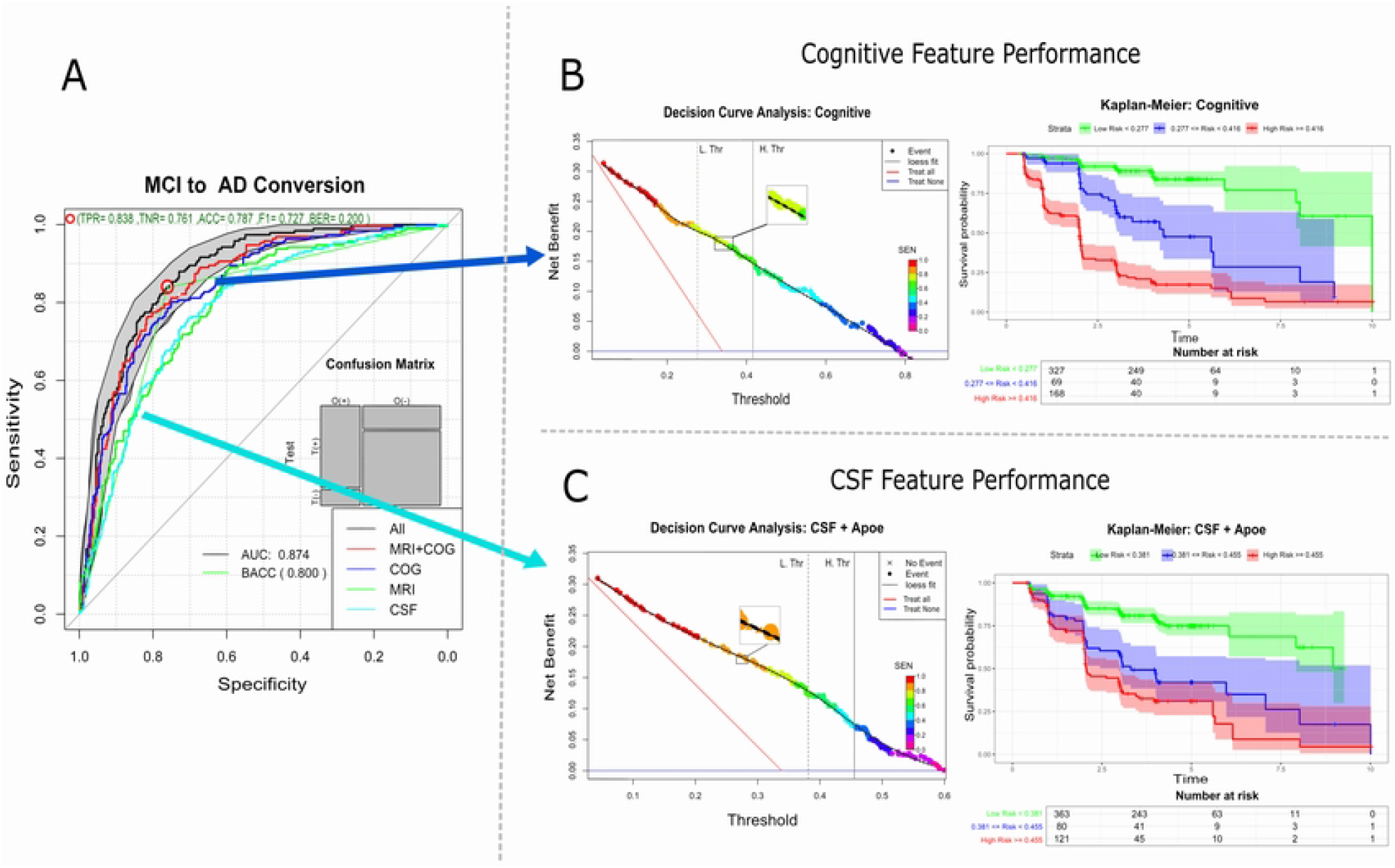
ROC curves, DCA and Kaplan-Meier. **(A)** ROC curves of method comparison experiment. **(B, C)** Plots shows the DCA and Kaplan-Meier performance on cognitive and CSF feature performance.

### Biomarker evaluation

To investigate the specific biomarkers contributing to the improved risk prediction, we analyzed the coefficients of the Cox proportional hazards models. Table 4 shows the beta coefficients and their corresponding hazard ratios with their corresponding 95% confidence intervals of the features that statistically improved the Cox model for the accurate high-risk, low-risk classification task. The table also shows that all data sources have features that aid the classification. The top feature was the Rey Auditory Verbal Learning Test, that indicated that higher scores indicate subjects that will remain stable with a z-standardized HR of 0.527 95% CI [0.400, 0.695]. The top CSF biomarker was the amyloid-*β* (A*β*), HR 0.679 95% CI [0.541, 0.853]; the *APOE ε*4 showed an HR 2.063 95% CI [1.357, 3.136]. The top qMRI feature was the mean volume of left entorhinal gray matter, whose HR was 0.689 95% CI [0.557, 0.852].

**Table 4.**
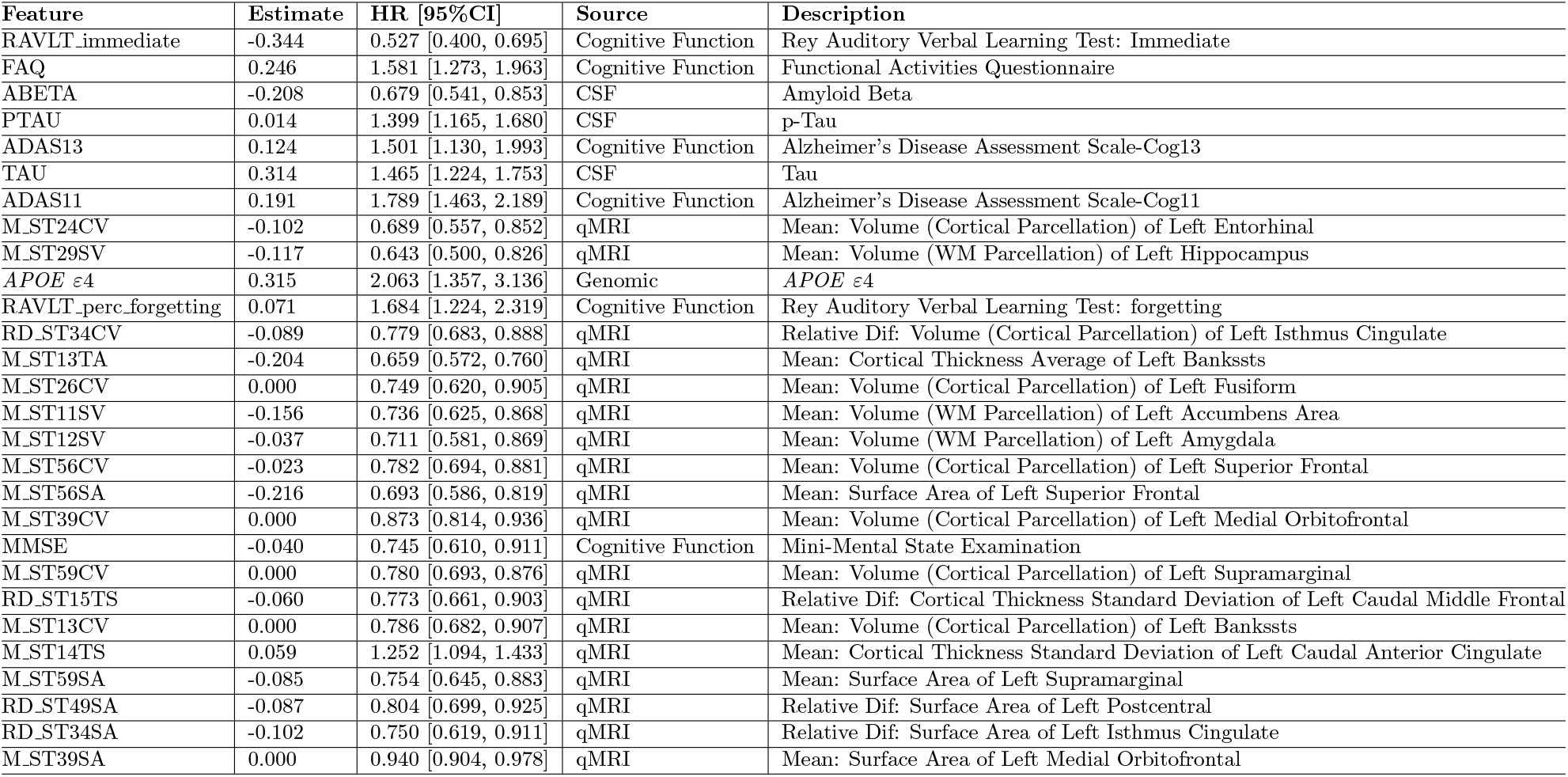
Top Features characteristics that intervene in the conversion of MCI to Alzheimer like relevant biomarkers.

The top left heatmap of Fig 5 shows the heatmap of the top features and depicts the features positively associated with conversion and the features with negative association to conversion. The graph at the bottom left of Fig 5 shows the association of variables in predicting conversion. Two main clusters of variables were found: the first cluster includes amyloid-*β* (A*β*), FAQ, and ADAS13, while the second cluster involves tau (p-tau), *APOE ε*4, and ADAS11. The plots on the left side report the performance of the model on the testing set. The ROC curve, the DCA and the Kaplan-Meier show that the model created by standardized features had very similar performance as predicted by the RHOCV procedure.

**Fig 5.**
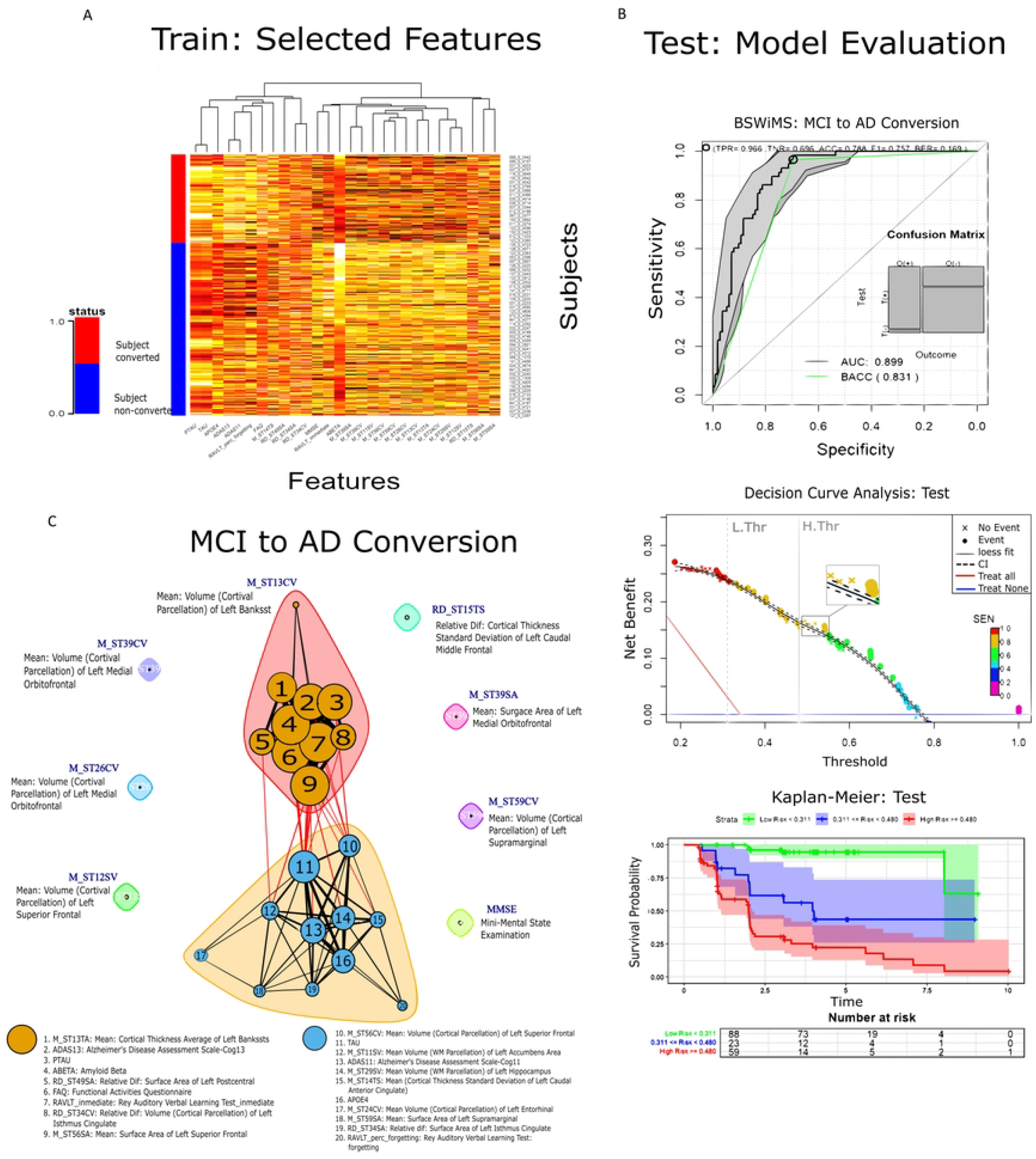
Feature Selection and Model Evaluation for MCI to Dementia Conversion Prediction. **(A)** Heat map of selected features at training phase, features are classified from subject conversion or non conversion. **(B)** Evaluation of the predictions on the testing set. ROC, DCA and Kaplan-Meier plots. **(C)** Graph clusters from the different features used in training.

## Discussion

In this section, we explore the potential of machine learning techniques combined with Cox models, with the aim of predicting the conversion from MCI to AD using multimodal data from subjects with MCI. As mentioned in the introduction, combining biomarkers from CSF, cognitive assessments, MRI features, and *APOE ε*4 status allows for a more comprehensive understanding of the dynamics associated with disease progression.

### ML feature subset selection

We validated that the integration of multiple biomarker modalities—CSF, *APOE ε*4, MRI, and cognitive assessments in Cox models for predicting MCI-to-AD conversion—is effective. Our validation was based on a stringent hold-out cross-validation scheme that randomly repeated the model training 50 times and analyzed the test results by advanced performance analysis methods like decision curve analysis and Kaplan-Meier plots. This analysis indicated that machine learning models such as LASSO and BSWiMS demonstrated their capacity to capture complex biomarker interactions, improving diagnostic precision and enabling more personalized treatment strategies. Recent research [47], highlighted the utility of LASSO in AD prediction, particularly when used with multimodal data, supporting our findings. The model’s capability to integrate cognitive and imaging biomarkers makes it well-suited to capture subtle patterns indicative of early AD progression.

Similarly, the BSWiMS model also showed top performance, especially in its ability to be the best model to predict conversion at 90% sensitivity, yielding an AUC of 0.87 with a sensitivity of 0.69 95% CI [0.63, 0.76]. Furthermore, BSWiMS showed a strong net benefit in a threshold range of 0.2-0.7, further reinforcing its reliability in clinical applications. As noted in a study [48], models that integrate structural MRI with other biomarkers, such as BSWiMS, tend to provide higher predictive accuracy, particularly for identifying patients at high risk of AD conversion. The ability of the BSWiMS model to stratify patients into distinct risk groups, as seen in Kaplan-Meier analysis, makes it a valuable tool for early diagnosis and risk management in clinical practice. Overall, the performance of both LASSO and BSWiMS models validates the importance of multimodal approaches for accurate and clinically relevant AD prediction.

### Source analysis and recent studies comparison

The study demonstrated differences in AUC values across the five experimental data-source conditions, highlighting the varying predictive power of different combinations of biomarkers. The integrated model with all biomarker modalities (CSF, *APOE ε*4, MRI, and cognitive data) achieved the highest AUC of 0.87, 95% CI [0.85, 0.90], indicating that multimodal integration significantly improves classification accuracy. This result is consistent with recent studies [47, 49], showing that the combination of multiple sources of biomarkers offers a more comprehensive understanding of disease pathology and leads to improved predictive performance. Conversely, models based on single modalities, such as CSF + *APOE ε*4, and MRI + *APOE ε*4, displayed a lower predictive power, suggesting that single-source biomarkers do not capture the full complexity of MCI-to-AD progression.

Comparing our methodology with a deep learning survival model, a study [50], worked with a deep learning-based survival model (DeepSurv) which included demographics, cognitive tests, genetic data, CSF biomarkers, and MRI measures as their features showed an accuracy of 0.83, while our best model BSWiMS, achieved similar results: 0.81, 95% CI [0.78, 0.84], with the added advantage that the BSWiMS Cox model is highly interpretable.

### Cox model feature analysis

Likewise, this study validated many of the well-established predictors of MCI-to-AD conversion. Notable examples [51, 52, 53], include p-Tau, Tau, *APOE ε*4, Amyloid Beta, the Functional Activities Questionnaire (FAQ), the Rey Auditory Verbal Learning Test (RAVLT), and specific brain regions such as the entorhinal and hippocampal cortices.

While our model aligns with recent studies in identifying several MRI features associated with MCI-to-AD conversion, it also highlights additional MRI features that emerged as significant, despite not being traditionally recognized as relevant indicators for this progression. A research study [52], identified the average cortical thickness in the superior frontal, medial orbitofrontal, caudal anterior cingulate, and isthmus cingulate regions as significant predictors. In contrast, our study highlights the significance of the mean volume in the cortical parcellation of the left superior frontal region, the mean surface area of the left medial orbitofrontal region, the mean cortical thickness standard deviation of the left caudal anterior cingulate, and relative difference on the volume from cortical parcellation on left isthmus cingulate as well as the mean surface area of the same region. These differences highlight the importance of looking beyond average cortical thickness, considering instead localized asymmetries and alternative measurements, such as surface area and mean volume within cortical parcellations.

In a broader context of feature relevance, a systematic review [54], identified that most studies prioritize MRI features, particularly entorhinal and hippocampal volumes, as key indicators for MCI-to-AD conversion. These features are also highlighted as critical components in our final model. Furthermore, the review indicates that many researchers combine structured data from multiple modalities. For instance, a study [55], used hippocampal volume as an MRI feature alongside A-beta as a CSF feature, while another study [56], included hippocampal volume, tau, and A-beta as features for predicting conversion. These studies align closely with our findings, as we incorporate both hippocampal and entorhinal volumes as features. Additionally, our model leverages multimodal data by integrating MRI, CSF, cognitive assessment, and genomic data, aligning with the multimodal approaches observed in related research.

## Data Availability

All relevant data are within the manuscript and its Supporting Information files.

## Limitations

The results presented in this work are limited to four key aspects. First, patient misdiagnosis is present, hence affecting feature selection and model building. The AD diagnosis used in this study is not definitive, but according to the NINCDS-ADRDA [57], it is the probable diagnosis of AD with probable errors in 10% to 15% of cases. Second, the presented findings were based on the ADNI cohort and measurements; therefore, it is biased toward the environmental factors present in the US and the Caucasian race. Third, qMRI results were based on FreeSurfer analysis; hence, changes in analytical tools may produce different results. Fourth, limited research has been done regarding the comparison of analytical techniques for automatic prediction of time to event within AD [58]. These key limitations indicate that the presented findings must be confirmed on cohorts from different countries and ethnicities.

## Conclusion

This study showed that multimodal biomarker integration and machine learning methods for the construction of interpretable Cox survival models are effective strategies in predicting the progression of mild cognitive impairment to Alzheimer’s disease. We improved diagnostic accuracy significantly by combining CSF fluid, *APOE ε*4 genotype, MRI, and cognitive tests over single-modality models. In particular, the LASSO and BSWiMS models performed admirably in identifying complicated biomarker relationships and stratifying patients into separate risk categories. Notably, the discovery of unique MRI features, such as the standard deviation of cortical thickness, emphasizes the necessity of investigating alternative MRI metrics other than usual averages. Our findings are consistent with previous research that has highlighted the usefulness of multimodal approaches and specific biomarkers such as p-Tau and hippocampal atrophy. Overall, this study provides strong evidence that our proposed methodology is clinically effective in early Alzheimer’s disease diagnosis and risk management. Furthermore, the discovery of novel MRI characteristics indicates the possibility of improving diagnostic criteria and establishing more targeted therapy strategies. Future studies should validate these findings in larger, more diverse populations and investigate the possibility of adding additional biomarkers, such as genetic and epigenetic variables. Finally, the findings from this study may help to design more accurate and tailored diagnostic methods for Alzheimer’s disease.

## Acknowledgments

This work was partially supported by Secretaría de Educación Superior, Ciencia, Tecnología e Innovación part of Gobierno de la República del Ecuador and by Strategic Research Group of Bioinformatics for Clinical Diagnosis from Tecnólogico de Monterrey. Data collection and sharing for this project was funded by the Alzheimer’s Disease Neuroimaging Initiative (ADNI) (National Institutes of Health Grant U01 AG024904) and DOD ADNI (Department of Defense award number W81XWH-12-2-0012). ADNI is funded by the National Institute on Aging, the National Institute of Biomedical Imaging and Bioengineering, and through generous contributions from the following: AbbVie, Alzheimer’s Association; Alzheimer’s Drug Discovery Foundation; Araclon Biotech; BioClinica, Inc.; Biogen; Bristol-Myers Squibb Company; CereSpir, Inc.; Cogstate; Eisai Inc.; Elan Pharmaceuticals, Inc.; Eli Lilly and Company; EuroImmun; F. Hoffmann-La Roche Ltd and its affiliated company Genentech, Inc.; Fujirebio; GE Healthcare; IXICO Ltd.; Janssen Alzheimer Immunotherapy Research & Development, LLC.; Johnson & Johnson Pharmaceutical Research & Development LLC.; Lumosity; Lundbeck; Merck & Co., Inc.; Meso Scale Diagnostics, LLC.; NeuroRx Research; Neurotrack Technologies; Novartis Pharmaceuticals Corporation; Pfizer Inc.; Piramal Imaging; Servier; Takeda Pharmaceutical Company; and Transition Therapeutics. The Canadian Institutes of Health Research is providing funds to support ADNI clinical sites in Canada. Private sector contributions are facilitated by the Foundation for the National Institutes of Health (www.fnih.org). The grantee organization is the Northern California Institute for Research and Education, and the study is coordinated by the Alzheimer’s Therapeutic Research Institute at the University of Southern California. ADNI data are disseminated by the Laboratory for Neuro Imaging at the University of Southern California.

## References

1. Association A. 2018 Alzheimer’s disease facts and figures. Alzheimer’s & Dementia. 2018;14(3):367–429.

2. Sperling RA, Aisen PS, Beckett LA, Bennett DA, Craft S, Fagan AM, et al. Toward defining the preclinical stages of Alzheimer’s disease: Recommendations from the National Institute on Aging-Alzheimer’s Association workgroups on diagnostic guidelines for Alzheimer’s disease. Alzheimer’s & Dementia. 2011;7(3):280–292.

3. Joachim CL, Morris JH, Selkoe DJ. Clinically diagnosed Alzheimer’s disease: Autopsy results in 150 cases. Annals of Neurology. 1988;24(1):50–56.

4. Salvatore C, Cerasa A, Castiglioni I. MRI Characterizes the Progressive Course of AD and Predicts Conversion to Alzheimer’s Dementia 24 Months Before Probable Diagnosis. Frontiers in Aging Neuroscience. 2018;10:135.

5. Lin SY, Lin PC, Lin YC, Lee YJ, Wang CY, Peng SW, et al. The Clinical Course of Early and Late Mild Cognitive Impairment. Frontiers in Neurology. 2022;13:685636.

6. Orozco-Sanchez J, Tamez-Peña JG. Prediction of MCI to AD Risk of Conversion Survival Models: qMRI vs CSF Measures and Cognitive Assessments. Medical Imaging. 2020;.

7. Jung NY, Kim ES, Kim HS, et al. Comparison of Diagnostic Performances Between Cerebrospinal Fluid Biomarkers and Amyloid PET in a Clinical Setting. Journal of Alzheimer’s Disease: JAD. 2020;.

8. Raghavan N, Samtani MN, Farnum M, Yang E, Novak G, Grundman M, et al. The ADAS-Cog revisited: Novel composite scales based on ADAS-Cog to improve efficiency in MCI and early AD trials. Alzheimer’s & Dementia. 2013;9(1 SUPPL.).

9. Corder EH, Saunders AM, Strittmatter WJ, Schmechel DE, Gaskell PC, Small GW, et al. Gene dose of apolipoprotein E type 4 allele and the risk of Alzheimer’s disease in late onset families. Science. 1993;.

10. Martínez-Torteya A, Treviño V, Tamez-Peña JG. Improved Diagnostic Multimodal Biomarkers for Alzheimer’s Disease and Mild Cognitive Impairment. Biomedical Research International. 2015; p. 1–11.

11. Shaffer JL, Petrella JR, Sheldon FC, Choudhury KR, Calhoun VD, Coleman RE, et al. Predicting Cognitive Decline in Subjects at Risk for Alzheimer Disease by Using Combined Cerebrospinal Fluid, MR Imaging, and PET Biomarkers. Radiology. 2013;266(2):583–591. doi:10.1148/radiol.12120010.

12. Paterson RW, Toombs J, Slattery CF, Nicholas JM, Andreasson U, Magdalinou NK, et al. Dissecting IWG-2 typical and atypical Alzheimer’s disease: insights from cerebrospinal fluid analysis. Journal of Neurology. 2015;262(12):2722–2730. doi:10.1007/s00415-015-7884-1.

13. Doherty CM, Forbes RB. Diagnostic lumbar puncture. Ulster Medical Journal. 2014;83(2):93–102.

14. Paterson RW, Slattery CF, Poole T, Nicholas JM, Magdalinou NK, Toombs J, et al. Cerebrospinal fluid in the differential diagnosis of Alzheimer’s disease: Clinical utility of an extended panel of biomarkers in a specialist cognitive clinic. Alzheimers Research and Therapy. 2018;10(1). doi:10.1186/s13195-018-0361-3.

15. Maddalena A, Papassotiropoulos A, Müller-Tillmanns B, Jung HH, Hegi T, Nitsch RM, et al. Biochemical diagnosis of Alzheimer disease by measuring the cerebrospinal fluid ratio of phosphorylated tau protein to beta-amyloid peptide42. Archives of Neurology. 2003;60(9):1202–1206.

16. Cullen B, O’Neill B, Evans JJ, Coen RF, Lawlor BA. A review of screening tests for cognitive impairment. Journal of Neurology, Neurosurgery and Psychiatry. 2007;78:790–799.

17. Kirsebom BE, Espenes R, Waterloo K, Hessen E, Johnsen SH, Bråthen G, et al. Screening for Alzheimer’s Disease: Cognitive Impairment in Self-Referred and Memory Clinic-Referred Patients. Journal of Alzheimer’s Disease. 2017;60(4):1621–1631.

18. Llano DA, Laforet G, Devanarayan V. Derivation of a new ADAS-cog composite using tree-based multivariate analysis: prediction of conversion from mild cognitive impairment to Alzheimer disease. Alzheimer Disease and Associated Disorders. 2013;25(1):73–84.

19. Da X, Toledo JB, Zee J, Wolk DA, Xie SX, Ou Y, et al. Integration and relative value of biomarkers for prediction of MCI to AD progression: spatial patterns of brain atrophy, cognitive scores, APOE genotype and CSF biomarkers. NeuroImage: Clinical. 2014;4:164–173.

20. Smith JJ, Sorensen AG, Thrall JH. Biomarkers in Imaging: Realizing Radiology’s Future. Radiology. 2003;227(3):633–638.

21. McEvoy LK, Brewer JB. Quantitative structural MRI for early detection of Alzheimer’s disease. Expert Review of Neurotherapeutics. 2010;10(11):1675–1688.

22. Fox NC, Schott JM. Imaging cerebral atrophy: normal ageing to Alzheimer’s disease. The Lancet. 2004;363(9406):392–394.

23. Mashal Y, Abdelhady HG, Iyer AK. Comparison of Tau and Amyloid-Targeted Immunotherapy Nanoparticles for Alzheimer’s Disease. Biomolecules. 2022;12(1):1–15. doi:10.3390/biom12010015.

24. Celaya-Padilla JM, Galván-Tejada CE, López-Monteagudo FE, Alonso-González O, Moreno-Báez A, Martínez-Torteya A, et al. Speed bump detection using accelerometric features: A genetic algorithm approach. Sensors (Switzerland). 2018;18(2):1–10.

25. Sarica A, Aracri F, Bianco MG, Arcuri F, Quattrone A, Quattrone A. Explainability of random survival forests in predicting conversion risk from mild cognitive impairment to Alzheimer’s disease. Brain Informatics. 2023;10(1):1–17. doi:10.1186/s40708-023-00211-w.

26. Liu K, Chen K, Yao L, Guo X. Prediction of Mild Cognitive Impairment Conversion Using a Combination of Independent Component Analysis and the Cox Model. Frontiers in Human Neuroscience. 2017;11:33. doi:10.3389/fnhum.2017.00033.

27. Zeifman LE, Eddy WF, Lopez OL, Kuller LH, Raji C, Thompson PM, et al. Voxel Level Survival Analysis of Grey Matter Volume and Incident Mild Cognitive Impairment or Alzheimer’s Disease. Journal of Alzheimer’s Disease. 2015;46(1):167–178. doi:10.3233/JAD-150089.

28. Michaud TL, Su D, Siahpush M, Murman DL. The Risk of Incident Mild Cognitive Impairment and Progression to Dementia Considering Mild Cognitive Impairment Subtypes. Dementia and Geriatric Cognitive Disorders Extra. 2017;7(1):15–29. doi:10.1159/000456633.

29. Tamez-Pena J, Martinez-Torteya A, Alanis I. Package ‘FRESA.CAD’ Feature Selection Algorithms for Computer Aided Diagnosis. CRAN Repository. 2016;.

30. Bichindaritz I, Englebert C, Regua A, Kotula L. Feature Selection and Case-Based Reasoning for Survival Analysis in Bioinformatics. The Thirty-First International Flairs Conference. 2018;.

31. Aguirre-Gamboa R, Martinez-Ledesma E, Gomez-Rueda H, Palacios R, Fuentes-Hernandez I, Sánchez-Canales E, et al. Efficient Gene Selection for Cancer Prognostic Biomarkers Using Swarm Optimization and Survival Analysis. Current Bioinformatics. 2016;11(3):310–323. doi:10.2174/1574893611666160615135456.

32. Schwarz G. Estimating the Dimension of a Model. The Annals of Statistics. 1978;6(2):461–464. doi:10.1214/aos/1176344136.

33. Marinescu RV, Oxtoby NP, Young AL, Bron EE, Toga AW, Weiner MW, et al. TADPOLE Challenge: Prediction of Longitudinal Evolution in Alzheimer’s Disease. The TADPOLE Challenge. 2018;.

34. Morris JC. The Clinical Dementia Rating (CDR): Current Version and Scoring Rules. Neurology. 1993;43(11):2412–2414. doi:10.1212/WNL.43.11.2412.

35. Rosen WG, Mohs RC, Davis KL. A new rating scale for Alzheimer’s disease. American Journal of Psychiatry. 1984;141(11):1356–1364.

36. Folstein M, Folstein S, Folstein J. The Mini-Mental State Examination: A Brief Cognitive Assessment. In: Principles and Practice of Geriatric Psychiatry: Third Edition. John Wiley and Sons; 2010. p. 145–146.

37. Bean J. Rey Auditory Verbal Learning Test, Rey AVLT. In: Encyclopedia of Clinical Neuropsychology. New York, NY: Springer New York; 2011. p. 2174–2175.

38. Moradi E, Hallikainen I, Hänninen T, Tohka J, Initiative ADN. Rey’s Auditory Verbal Learning Test scores can be predicted from whole brain MRI in Alzheimer’s disease. Neuroimage Clin. 2017;13:415–427. doi:10.1016/j.nicl.2016.12.018.

39. Collett D. Modelling Survival Data in Medical Research. Chapman & Hall/CRC; 2003.

40. Tamez-Pena J. Feature Selection and the BSWiMS Method; 2018.

41. Tibshirani R. Regression Shrinkage and Selection via the Lasso. Journal of the Royal Statistical Society Series B (Methodological). 1996;.

42. Wen C, Zhang A, Quan S, Wang X. BeSS: An R Package for Best Subset Selection in Linear, Logistic and CoxPH Models; 2017.

43. Simon N, Friedman J, Hastie T, Tibshirani R. Regularization Paths for Cox’s Proportional Hazards Model via Coordinate Descent. Journal of Statistical Software. 2011;.

44. Robin X, Turck N, Hainard A, Tiberti N, Lisacek F, Sanchez JC, et al. pROC: an open-source package for R and S+ to analyze and compare ROC curves. BMC Bioinformatics. 2011;12:77. doi:10.1186/1471-2105-12-77.

45. Kassambara A, Kosinski M, Biecek P. Package ‘survminer’: Drawing Survival Curves using ‘ggplot2’; 2017. Available from: https://CRAN.R-project.org/package=survminer.

46. Mantel N. Evaluation of survival data and two new rank order statistics arising in its consideration. Cancer Chemotherapy Reports. 1966;50(3):163–170.

47. Liu M, Cheng D, Wang X, et al. Multimodal Neuroimaging Feature Selection with Consistent Metric Constraint for Diagnosis of Alzheimer’s Disease. Medical Image Analysis. 2020;60:101625. doi:10.1016/j.media.2020.101625.

48. Mito R, Li Q, Doecke J, et al. Combining MRI and PET Biomarkers Improves Prediction of Progression from Mild Cognitive Impairment to Alzheimer’s Disease. Alzheimer’s Research Therapy. 2021;13:99. doi:10.1186/s13195-021-00804-7.

49. Leuzy A, Mattsson-Carlgren N, Palmqvist S, et al. Biomarker-Based Prediction of Progression in Preclinical Alzheimer’s Disease. Alzheimer’s Dementia. 2022;18:573–583. doi:10.1002/alz.12694.

50. Mirabnahrazam G, Ma D, Beaulac C, et al. Predicting Time-to-Conversion for Dementia of Alzheimer’s Type Using Multi-Modal Deep Survival Analysis. Neurobiology of Aging. 2023;121:139–156. doi:10.1016/j.neurobiolaging.2023.07.015.

51. Delli Pizzi S, Punzi M, Sensi SL. Functional Signature of Conversion of Patients with Mild Cognitive Impairment. Neurobiology of Aging. 2019;74:21–37. doi:10.1016/j.neurobiolaging.2018.11.012.

52. Varatharajah Y, Ramanan VK, Iyer R, Vemuri P, Initiative ADN. Predicting Short-Term MCI-to-AD Progression Using Imaging, CSF, Genetic Factors, Cognitive Resilience, and Demographics. Scientific Reports. 2019;9:2235. doi:10.1038/s41598-019-38699-9.

53. Chen Y, Qian X, Zhang Y, et al. Prediction Models for Conversion from Mild Cognitive Impairment to Alzheimer’s Disease: A Systematic Review and Meta-Analysis. Frontiers in Aging Neuroscience. 2022;14:496. doi:10.3389/fnagi.2022.00326.

54. Muhammed Niyas KP, Thiyagarajan P. A Systematic Review on Early Prediction of Mild Cognitive Impairment to Alzheimer’s Using Machine Learning Algorithms. International Journal of Intelligent Networks. 2023;4:74–88. doi:10.1016/j.ijin.2023.05.003.

55. Lei B, Yang P, Wang T, Chen S, Ni D. Relational-regularized discriminative sparse learning for Alzheimer’s disease diagnosis. IEEE Transactions on Cybernetics. 2017;47(4):1102–1113.

56. Frölich L, Peters O, Lewczuk P, Gruber O, Teipel SJ, Gertz HJ, et al. Incremental value of biomarker combinations to predict progression of mild cognitive impairment to Alzheimer’s dementia. Alzheimer’s Research Therapy. 2017;9:1–15.

57. Thal LJ, Kantarci K, Reiman EM, Klunk WE, Weiner MW, Zetterberg H, et al. The role of biomarkers in clinical trials for Alzheimer disease. Alzheimer Disease & Associated Disorders. 2006;20(1):6–15.

58. Billichová M, Coan LJ, Czanner S, Kováčová M, Sharifian F, Czanner G. Comparing the performance of statistical, machine learning, and deep learning algorithms to predict time-to-event: A simulation study for conversion to mild cognitive impairment. Plos one. 2024;19(1):e0297190.

